# Perioperative antimicrobial prophylaxis in patients receiving antibiotic therapy

**DOI:** 10.1101/2021.06.14.21255125

**Authors:** Yiwei Yin, Eljim P Tesoro, Alan E Gross, Jeffrey J Mucksavage

## Abstract

**Objective:** Antimicrobial prophylaxis is administered perioperatively to prevent surgical site infections. However, in patients who have already received antibiotics for the treatment of active infections prior to surgery, the risks and benefits of administering prophylactic antibiotics are unknown. We aimed to assess the necessity of perioperative prophylactic antibiotic administration in patients receiving antibiotic treatment for active infections.

**Method:** This was a retrospective, chart-review cohort study. Between January 2018 to May 2018, adult patients who underwent inpatient surgery at the University of Illinois Hospital and Health Sciences System, and were prescribed prophylactic antibiotics based on institutional protocol, while receiving antibiotic treatment within 48 hours prior to surgery, were included in the study. The primary endpoint was the rate of duplicative antibiotic therapy, which was defined as the administered prophylactic antibiotic (1) exhibiting similar or narrower bacterial coverage as the treatment antibiotic(s), and (2) given within the dose interval of the treatment antibiotic(s).

**Results:** A total of 158 patients were included in the study, of which 70 (44.3%) received duplicative antibiotic therapy, whereas 88 (55.7%) did not. Differences in the incidence of acute kidney injury, *C. difficile* infection, and surgery site infections were not statistically significant between the two groups.

**Conclusion:** We found that it was common for patients receiving therapeutic systematic antibiotics to perioperatively be prescribed additional prophylactic antibiotics at our institution. However, additional prophylactic antibiotics can be unnecessary in decreasing the incidence of surgical site infections but may increase the risk of adverse reaction.

## Introduction

Prophylactic antibiotics are administered perioperatively to prevent infections at the surgical site. The selection and administration of antibiotics is based on the likely infecting organism(s), any active infection, patient characteristics, likelihood of bacterial resistance, and wound classification (i.e., dirty vs. clean) [1]. Based on the recommendations from the American Society of Health-System Pharmacist (ASHP) and the Society for Healthcare Epidemiology of America clinical practice guidelines, the administration of prophylactic antibiotics is usually started within 60 minutes of surgery and stopped no longer than 24 hours after surgery [1,2]. Multiple studies have shown that the timing of perioperative prophylactic antibiotic administration is critical for the successful prevention of surgical site infections (SSI). Administration of these antibiotics in close proximity to the site of incision helps ensure that adequate serum and tissue concentrations are achieved for adequate antimicrobial [1–3].

In practice, patients treated for active infections prior to surgery may end up receiving antibiotics for both treatment and prophylaxis, resulting in duplicative antibiotic therapy (DAT). The ASHP guidelines recommend the administration of prophylactic antibiotics in patients receiving antibiotics for the treatment of remote infections. However, this recommendation is based solely on the intention of ensuring adequate tissue concentration of the antimicrobial agent and is not supported by clinical studies or pharmacokinetic data. Moreover, the ASHP guideline does not discuss the risks involved in administering prophylactic antibiotics to these patients; therefore, evidence is limited. A small, retrospective study conducted at the University of Chicago found that DAT provided no additional benefit in preventing SSIs. In addition, this study noted a rise in antibiotic-associated side effects, including infusion reaction and nephrotoxicity, in patients receiving DAT [1]. The actual risks and benefits of DAT are unclear and have not been explored extensively. It is still unclear whether DAT can play a role in reducing surgical site infections. Moreover, DAT can increase the risk of postoperative acute kidney injury (AKI) and *Clostridium difficile* infection, as well as the cost of treatment. It may also increase the emergence of microbial resistance. Therefore, further research is required to gain a better understanding of the potential risks and benefits of DAT.

## Methods

### Study Design

This was a retrospective, descriptive chart review study involving adult patients (<u>≥</u> 18 years) who had undergone inpatient surgery at the University of Illinois Hospital & Health Science System (UIH) between 1 January 2018 and 8 May 2018. The study was approved by the Institutional Revie Board (IRB) of University of Illinois at Chicago, with a waiver of informed consent. Patients were identified from operating room records using electronic medical record (EMR) system (Cerner [Cerner lnc., Kansas City, Missouri]). Patients were included if they required antibiotic prophylaxis per the UIH antimicrobial surgical prophylaxis protocol and had received other antibiotics for the treatment of active infection within 48 hours before the surgery. Patients were excluded if they underwent surgery within 24 hours of transferring from another institution or home, if they underwent outpatient surgery, or if they were pregnant. The appropriateness of the choice of prophylactic antibiotic based on the type of surgery was not assessed; all patients included in the study had suspected or confirmed infection, so the choice of prophylactic antibiotic may have taken active infection into consideration. The primary endpoint of this study was the rate of DAT. DAT was defined as the prescription of prophylactic antibiotic (1) with similar or narrower bacterial coverage as the treatment antibiotic(s), and (2) administered within the dose interval of the treatment antibiotic(s). The secondary outcomes were compared between patients who did and did not receive DAT, including the incidence of postoperative AKI (defined as serum creatinine (Scr) over 1.5-fold of the baseline level within 48 hours post-procedure; excluding patients with end-stage renal disease on dialysis), documented incidence of *C. difficile* infection occurring within 30 days after the procedure, and documented SSIs reported within 30 days after the procedure.

### Data collection

Demographic data collected included weight, race, age, sex, allergies, type of surgery, type of infection, and wound classification. Wound classification was based on the National Health Care Safety Network criteria of the Centres for Disease Control and Prevention. Records of the antibiotic regimens for prophylaxis and treatment were obtained, along with the time of administration, immediately before or after surgery. Documentation of SSIs occurring within 30 days after the surgery was done to assess clinical outcomes. The incidence of postoperative AKI, Scr levels, and creatinine clearance were reviewed at the baseline (lowest value within 48 hours perioperative) and within 48 hours postoperatively (as available). A postoperative Scr level of over 1.5-fold the baseline was considered AKI. To assess other risk factors that may contribute to AKI, records of the occurrence of hypotension during surgery (defined as requiring pressor during surgery or systolic blood pressure < 90 mmHg), and the use of contrast within 48 hours before surgery were also collected. The incidence of *C. difficile* infection within 30 days after surgery was also considered to evaluate the safety of the treatment regimen.

### Statistical analysis

Patients were divided into two groups: the non-DAT group, comprising patients who did not receive DAT; and the DAT group, comprising patients who received DAT. Data was analysed for all the patients in both groups. Descriptive statistics were used to analyse demographic and clinical characteristics. For categorical data, Fisher’s exact test or chi-square test was used, depending on the sample size. Student’s *t*-test was used to analyse continuous data that exhibited an approximately normal distribution, while the Mann Whitney U test was used for continuous data that was not normally distributed. Differences were considered statistically significant at p < 0.05.

## Results

A total of 158 patients who underwent inpatient surgery between 1 January 2018 and 8 May 2018 met the inclusion criteria. Of these, 70 patients (44.3%) received DAT, and 88 (55.7%) were not administered prophylactic antibiotics in addition to the treatment antibiotics. All patients who received DAT were given prophylactic antibiotics with a similar or narrower spectrum of activity as the treatment antibiotics. Notably, 22 patients (31.4%) in the DAT group received the same antibiotics for both prophylaxis and treatment. In the non-DAT group, 59 patients (67.0%) received adequate antibiotics for the treatment of infection and did not receive additional prophylactic therapy. The remaining 29 patients (33.0%) received prophylactic antibiotics for surgery without duplication.

The baseline characteristics of patients in the DAT and non-DAT groups were similar (Table 1).

**Table 1:** Demographic characteristics, surgery and infection details.

|  | **All patients**<br>**N=158** | **Non-DAT**<br>**Group**<br>**N=88** | **With DAT**<br>**N=70** | **P-value** |
| --- | --- | --- | --- | --- |
| Age (years, Mean ± S.D.) | 55.7 ± 14.4 | 56.1 ± 14.5 | 55.2 ± 14.4 | 0.85 |
| Weight (kg, Mean ± S.D.) | 80.1 ± 23.6 | 78.1 ± 21.7 | 82.7 ± 25.9 | 0.28 |
| Male, no. (%) | 91 (57.6) | 54 (61.3) | 37 (52.9) | 0.37 |
| Any antibiotic allergy, no. (%) | 31 (19.6) | 22 (25) | 9 (13.6) | 0.45 |
| Race, no. (%)<sup>a</sup> |  |  |  | 0.56 |
| African American | 62 (39.2) | 35 (40.0) | 27 (38.6) |  |
| Asian | 2 (1.26) | 0 | 2 (2.9) |  |
| Caucasian | 38 (24.1) | 20 (22.7) | 18 (25.7) |  |
| Hispanic | 3 (1.9) | 2 (2.3) | 1 (1.4) |  |
| Other | 53 (33.5) | 31 (35.2) | 22 (31.4) |  |
| Infection type no. (%)<sup>b</sup> |  |  |  | 0.24 |
| Blood | 16 (10.1) | 10 (11.4) | 6 (8.5) |  |
| Bone | 21 (13.3) | 14 (15.9) | 7 (10.0) |  |
| CNS | 3 (1.9) | 1 (1.1) | 2 (2.9) |  |
| ENNT | 1 (0.6) | 0 | 1 (1.4) |  |
| Intrabdominal | 14 (8.9) | 9 (10.2) | 5 (7.1) |  |
| Lung | 12 (7.6) | 5 (5.7) | 7 (10.0) |  |
| Neutropenic fever | 1 (0.6) | 1 (1.1) | 0 |  |
| Skin/soft tissue | 35 (22.1) | 24 (27.3) | 11 (16.7) |  |
| Urine | 9 (5.7) | 5 (5.7) | 4 (5.7) |  |
| Wound | 28 (17.7) | 10 (11.4) | 18 (25.7) |  |
| Unknown | 29 (18.4) | 18 (20.5) | 11 (15.7) |  |
| Wound Classification, no. (%) |  |  |  | 0.84 |
| Clean-contaminated | 41 (25.9) | 23 (26.1) | 18 (25.7) |  |
| Contaminated | 22 (13.9) | 11 (12.5) | 11 (15.7) |  |
| Dirty-infected | 95 (60.1) | 54 (61.4) | 41 (58.5) |  |
| Surgery type, no. (%) |  |  |  | 0.14 |
| Cardiac | 2 (1.3) | 0 | 2 (2.9) |  |
| Colorectal | 2 (1.3) | 1 (1.1) | 1 (1.4) |  |
| Otorhinolaryngology | 2 (1.3) | 0 | 2 (2.9) |  |
| Gastrointestinal | 41 (25.9) | 22 (25.0) | 19 (27.1) |  |
| Head and neck | 4 (2.5) | 4 (4.5) | 0 |  |
| Neurosurgery | 10 (6.3) | 5 (5.7) | 5 (7.1) |  |
| Orthopedic | 25 (15.8) | 11 (12.5) | 14 (20.0) |  |
| Plastic | 27 (17.1) | 14 (15.9) | 13 (18.5) |  |
| Thoracic | 3 (1.9) | 1 (1.1) | 2 (2.9) |  |
| Urologic | 4 (2.5) | 3 (3.4) | 1 (1.4) |  |
| Vascular | 38 (24.1) | 27 (30.6) | 11 (15.7) |  |

<sup>a</sup>No specific detail was provided in patients with “other” race in electronic medical record
<sup>b</sup>Total of 11 patients was treated for two different infection at the same time, with 9 patients in DAT group and 2 patients in non-DAT group.

**Table 2:** Incidence of surgical site infection, *C. difficile* infection and postoperative acute kidney injury.

|  | **All**<br>**patients**<br>**N=158** | **Non-DAT**<br>**Group**<br>**N=88** | **DAT group**<br>**N=70** | **P-value** |
| --- | --- | --- | --- | --- |
| Surgical site infection within 30 days after surgery, no. (%) | 3 (1.9) | 2 (2.1) | 1(1.4) | >0.99 |
| *C. difficile* infection within 30 days after surgery, no. (%) | 1 (0.6) | 0 | 1 (1.4) | 0.44 |
| Postoperative acute kidney injury within 48 hours after surgery, no. (%) | 14 (5.6) | 5 (5.3) | 9 (12.9) | 0.16 |
| Hypotension | 4 | 2 | 2 |  |
| Contrast use within 48 hours before the surgery | 1 | 0 | 1 |  |

Total of 14 patients experienced AKI, of which five patients were in the appropriate antibiotic use group, while nine patients were in the inappropriate use group. AKI in three patients could be attributed to hypotension during surgery or use of contrast prior to surgery. The differences in the secondary outcomes were not statistically significant. In the DAT group (Table 3), five patients received additional vancomycin, and notably, three patients developed AKI after surgery.

**Table 3:** Prophylactic antibiotic used in patient received DAT.

| Prophylaxis antibiotics | Number of patients<br>N= 70 |
| --- | --- |
| Cefazolin | 38 |
| Ceftriaxone | 7 |
| Piperacillin/tazobactam | 6 |
| Vancomycin | 5 |
| Clindamycin | 4 |
| Metronidazole | 4 |
| Ampicillin/Sulbactam | 3 |
| Imipenem or Imipenem/Cilastatin | 3 |
| Levofloxacin | 1 |
| Cefepime | 1 |

## Discussion

This retrospective study evaluated DAT in patients undergoing surgery, who were administered antibiotics for the treatment of infection, as well as for surgical prophylaxis. The study revealed opportunities for stewardship and quality improvement. Nearly half of the patients (44.3%) received DAT, and 31.4% received the same antibiotics for prophylaxis and treatment in the DAT group. Although DAT was not found to be associated with a statistically significant increase in adverse events in this study, there was a numerical increase in the percentage of postoperative cases of AKI in the DAT group (12.9% and 5.7% in the DAT and non-DAT group, respectively). Three patients in the DAT group developed AKI after receiving vancomycin for both treatment and prophylaxis, resulting in a supratherapeutic postoperative vancomycin serum concentration (>20 µg/mL). This raises the concern of increasing the risk of AKI with duplicated antibiotic use. This study did not find a decrease in the incidence of SSIs with DAT. Therefore, the benefits of administering additional prophylactic antibiotics prior to surgery, while patients already have adequate antibiotic coverage are questionable.

There are limited guidelines regarding the perioperative use of prophylactic antimicrobials in patients being treated for systemic infections. The guidelines published by multiple organisations, including the Society of Thoracic Surgeons (STS), American College of Obstetricians and Gynaecologists, and Centres for Disease Control and Prevention, and American Urological Association (AUA), do not address antimicrobial prophylaxis in patients with active infections [4–7]. The AUA and STS guidelines exclude this population altogether, since it is not within the focus of the guidelines [6,7]. ASHP, on the other hand, offers a specific recommendation, supporting the use of perioperative antimicrobial prophylaxis in patients being treated for an active infection to ensure adequate tissue concentration. However, there is limited evidence to support this recommendation [8]. Several studies have shown that tissue or serum antibiotic concentrations exceeding the minimal inhibitory concentration of the targeted microorganism are important for the effective prevention of SSIs [9–12]. However, none of these studies included patients who were being treated for active infections. A retrospective study including 499 patients undergoing surgery evaluated the effect of administration of additional prophylactic antibiotics in patients on antibiotic treatment. Similar to our study, the authors observed no difference in the incidence of SSIs but did see an increase in antibiotic-related side effects. In our study, 48 patients (68.6%) in the DAT group received at least three doses of systematic antibiotics at regular intervals within 48 h before the surgery. The benefits of DAT may be limited since the patients in our study had already received adequate doses of antibiotics.

This study had several limitations. Firstly, the sample size was limited, making it difficult to identify differences in secondary outcomes. Secondly, in a retrospective study, the quality of data depends on the quality of documentation in the EMRs, and therefore, information may be missing due to a lack of follow-up or inadequate documentation. Additionally, this study did not have a comprehensive review of the confounders for secondary outcomes (e.g., AKI). Moreover, since the patients had active infections, the choice of prophylactic antibiotics could be complicated and may not have followed general guidelines or hospital protocols. Therefore, we did not evaluate the appropriateness of the choice of prophylactic antibiotic.

As one of the first few studies to evaluate this specific clinical scenario, this study showed that DAT may not decrease the incidence of SSIs but could increase the risk of associated adverse effects. Prospective research is required to investigate whether additional prophylactic antibiotics are necessary in patients receiving antibiotic treatment or reaching a steady state. We still lack information on the incidence of SSIs, and its correlation with prophylactic antibiotic use in this population, since this has not yet been studied. Further studies are warranted to provide a comprehensive risk-benefit analysis of DAT, taking into consideration the cost to treatment, as well as the risk of antimicrobial resistance. Generally, perioperative antibiotic use is not reviewed while conducting prophylactic antibiotic assessment. Our findings also reveal great opportunities for antimicrobial stewardship, to improve appropriate prescription of prophylactic antibiotics in patients with active infections. Different approaches, such as updating systematic guidelines and developing appropriate system alerts, may be applied. Pharmacists can play an important role in reviewing the history of antibiotic use, conducting patient-specific antibiotic assessments, and increasing team awareness of potential DAT.

## Conclusion

Our study found that it was common for patients receiving therapeutic systematic antibiotics to be prescribed additional prophylactic antibiotics perioperatively at our institution. However, additional prophylactic antibiotics can be unnecessary and may not be beneficial in decreasing the incidence of SSIs, but may increase the risk of adverse reaction.

## What this paper adds

### What is already known on this subject

- There is limited evidence regarding the use of perioperative prophylactic antimicrobials in patients being treated for systematic infections.
- The incidence for patients to receive both perioperative prophylactic antibiotics and treatment antibiotics is not well reported

### What this study adds

- It can be common for patients to receive both perioperative prophylactic antibiotics and treatment antibiotics
- No significant differences in the incidence of surgical site infections with duplicate antibiotic use, however, the incidence of antibiotic-related side effects, including acute kidney injury, may be increased.
- Our study reveals opportunities for stewardship and quality improvement in optimizing antibiotics use for patients undergoing surgery.

## Data Availability

N/A

## References

1. Dale B, E-Patchen D, Keith O, et al. Clinical practice guidelines for antimicrobial prophylaxis in surgery. Surg Infect (Larchmt) 2013;14(1):73–156.

2. Deverick A, Kelly P, Berríos-Torres S, et al. Strategies to prevent surgical site infections in acute care hospitals: 2014 update. Infect Control Hosp Epidemiol 2014;35(6):605–627.

3. James S, Barbara B, Walter H, et al. Timing of antimicrobial prophylaxis and the risk of surgical site infections: results from the trial to reduce antimicrobial prophylaxis errors. Ann Surg 2009; 250(1):10–6.

4. Major K, Judith M, Alewjin O, et al. Antibiotic prophylaxis and the risk of surgical site infections following total hip arthroplasty: timely administration is the most important factor. Clin Infect Dis 2007;44(7):921–7.

5. Joseph D, Joseph V, Talmadge B, et al. Intraoperative serum and tissue activity of cefazolin and cefoxitin. Arch Surg 1985;120(7):829–32.

6. C N, C R, T H, et al. Evaluation of surgical site infection antibiotic prophylaxis among patients receiving antibiotics for active infection. J Hosp Infect. 2019;103(3):354–5.

7. Archana A, Maggie S, Payal M, et al. Prolonged antimicrobial prophylaxis following cardiac device procedures increases preventable harm: insights from the VA CART program. Infect Control Hosp Epidemiol 2018;39(9):1030–6.

8. Westyn B, John R, William J, et al. Risk of surgical site infection, acute kidney injury, and Clostridium difficile infection following antibiotic prophylaxis with vancomycin plus a beta-lactam versus either drug alone: A national propensity-score-adjusted retrospective cohort study. PLoS Med. 2017;14(7): e1002340.

9. Ikemefuna O, Ramakanth Y, Lauren P, et al. Surgical Wound Classification and Surgical Site Infections in the Orthopaedic Patient. J Am Acad Orthop Surg Glob Res Rev. 2017;1(3):e022.

10. ACOG Practice Bulletin No. 195: Prevention of Infection After Gynecologic Procedures. Obstet Gynecol 2018 Jun;131(6): e172–e189.

11. S B, C U, D B, et al. Centers for Disease Control and Prevention Guideline for the Prevention of Surgical Site Infection 2017. JAMA Surg. 2017;152(8):784–91.

12. Fred E, Richard E, Peter H, et al. The Society of Thoracic Surgeons Practice Guideline Series: Antibiotic Prophylaxis in Cardiac Surgery, Part I: Duration Ann Thorac Surg. 2006;81(1):397–404.

13. Deborah L, Kevin W, Joyce S, et al Best Practice Statement on Urologic Procedures and Antimicrobial Prophylaxis. J Urol. 2020;203(2):351–6.

14. Walter W, Edin M, Marcel Z, et al. Timing of surgical antimicrobial prophylaxis: a phase 3 randomised controlled trial. Lancet Infect Dis. 2017;17(6):605–14.

